# How does the proportion of never treatment influence the success of mass drug administration programmes for the elimination of lymphatic filariasis?

**DOI:** 10.1101/2023.10.20.23297321

**Authors:** Klodeta Kura, Wilma A. Stolk, Maria-Gloria Basáñez, Benjamin S. Collyer, Sake J. de Vlas, Peter J Diggle, Katherine Gass, Matthew Graham, T. Déirdre Hollingsworth, Jonathan D. King, Alison Krentel, Roy M. Anderson, Luc E. Coffeng

## Abstract

**Background:** Mass drug administration (MDA) is the cornerstone for the elimination of lymphatic filariasis (LF). The proportion of the population that is never treated (NT) is a crucial determinant of whether this goal is achieved within reasonable timeframes.

**Methods:** Using two individual-based stochastic LF transmission models, we assess the maximum permissible level of NT for which the 1% mf prevalence threshold can be achieved (with 90% probability) within 10 years under different scenarios of annual MDA coverage, drug combination and transmission setting.

**Results:** For *Anopheles*-transmission settings, we find that treating 80% of the eligible population annually with ivermectin+albendazole (IA) can achieve the 1% mf prevalence threshold within 10 years of annual treatment when baseline mf prevalence is 10%, as long as NT <10%. Higher proportions of NT are acceptable when more efficacious treatment regimens are used. For *Culex*-transmission settings with a low (5%) baseline mf prevalence and Diethylcarbamazine+Albendazole (DA) or Ivermectin+Diethylcarbamazine+Albendazole (IDA) treatment, elimination can be reached if treatment coverage among eligibles is 80% or higher. For 10% baseline mf prevalence, the target can be achieved when the annual coverage is 80% and NT ≤15%. Higher infection prevalence or levels of NT would make achieving the target more difficult.

**Conclusions:** The proportion of people never treated in MDA programmes for LF can strongly influence the achievement of elimination and the impact of NT is greater in high transmission areas. This study provides a starting point for further development of criteria for the evaluation of NT.

## Introduction

Lymphatic filariasis (LF) is a mosquito-borne neglected tropical disease (NTD) caused by three parasites, namely, *Wuchereria bancrofti*, *Brugia malayi* and *Brugia timori* [1]. LF can cause chronic morbidity, such as hydrocele or lymphedema which are associated with disability, pain, mental health problems, reduced productivity, and social stigmatisation [2–4]. In 2000, the World Health Organization (WHO) established the Global Programme to Eliminate Lymphatic Filariasis (GPELF) with the target of eliminating the disease as a public health problem (EPHP) [5]. The two key goals of the programme are (i) interruption of transmission by using community-wide mass drug administration (MDA) for at least 5 years using a two-drug combination (ivermectin + albendazole [IA] in onchocerciasis co-endemic areas in Africa, and diethylcarbamazine citrate + albendazole [DA] elsewhere), and (ii) to reduce the suffering of patients by managing morbidity and preventing disability. In areas where *W. bancrofti* is endemic and *Anopheles* and/or *Culex* are the principal vectors, the first goal is considered to be met when the level of infection is reduced to less than 1% microfilaraemia (mf) prevalence in the population aged 5 years and above. The achievement of this goal is measured through a series of transmission assessment surveys (TAS) [6].

Great progress has been made towards the WHO target. By 2019, more than 8.6 billion treatments had been successfully distributed resulting in a 74% reduction in the number of individuals infected with LF [7]. WHO aims to validate elimination as a public health problem in 81% of endemic countries by 2030 [1]. To accelerate progress towards this goal in areas lagging behind, the WHO has recommended to use a combination of all three drugs, known as the triple drug (IDA) therapy in eligible settings [8,9]. Due to the risk of adverse events, this combination is not recommended in those African countries with LF-onchocerciasis or LF-loiasis co-endemic areas [8,9].

LF elimination programmes have been successful in some, but not all areas. An important determinant of reaching the 1% mf prevalence threshold is the population coverage of MDA programmes, which needs to be sufficiently high and is recommended to be at least 65% of the total population [5]. However, in addition to population coverage, prospects of achieving <1% mf prevalence have been recognised to depend on patterns regarding who does/does not take treatment in MDA programmes [10]. Especially in settings with persistent transmission after many rounds of MDA, the question arises as to whether there are groups of individuals who are sustaining transmission due to repeatedly missing treatment [15,16]. There are many reasons why someone may never take treatment, including intentional factors (i.e., refusal, non-attendance, failure to ingest, fear of side-effects) and unintentional factors (i.e., out of the village at the time of MDA, treatment not offered, not eligible for treatment) [11–14]. The term ‘never treated’ (NT) has been proposed to capture all causes for never treatment, irrespective of the reason or intentionality and refers to individuals who have never been treated across consecutive treatment rounds [15,16]. Other terms related to people not being treated (e.g., systematic non-compliance, non-participation, non-attendance) may refer to specific causes for non-treatment and those terms are not used in this work.

The achievement of the WHO 2030 goals may be hampered if too many people remain never treated [10,21,26,27]. Therefore, quantifying NT levels (and ultimately, understanding the reasons behind them so they can be minimised) is critically important. The programmatic implications of a particular level of NT are currently unclear. In this work, we provide modelling insight into the impact of NT on the likelihood that LF programmes achieve the 1% mf prevalence threshold in epidemiological settings where *Anopheles* or *Culex* mosquitoes are the main vectors.

## Methods

We use two individual-based stochastic models, namely, TRANSFIL and LYMFASIM to simulate the impact of NT on the probability of reaching the <1% mf prevalence threshold. Details of the two transmission dynamics models and their parameterization have been published previously [17–26,28].

### Models for never treatment patterns in MDA

There are many approaches to modelling patterns regarding the proportion of the population never treated [10,27]. In LYMFASIM, NT is the result of an input parameter for the proportion of people who will ultimately never be treated (dashed horizontal line in Figure 1) and the overall population coverage of MDA. Patterns in repeated (non-)treatment of eligible individuals are assumed to be the result of a mix of systematic and random factors, which is achieved by assigning simulated individuals a trait for their inclination to participate in MDA. This means that the proportion of eligible people that has never been treated at some time point is higher than expected under random treatment. However, the level of NT among eligibles is directly tied to the overall coverage level (such that for increasing MDA coverage there is a limit to the NT values that can be simulated). Therefore, in LYMFASIM, we simulate different levels of NT for a given MDA coverage by changing the parameter for the proportion of people who will never be treated.

**Figure 1:**
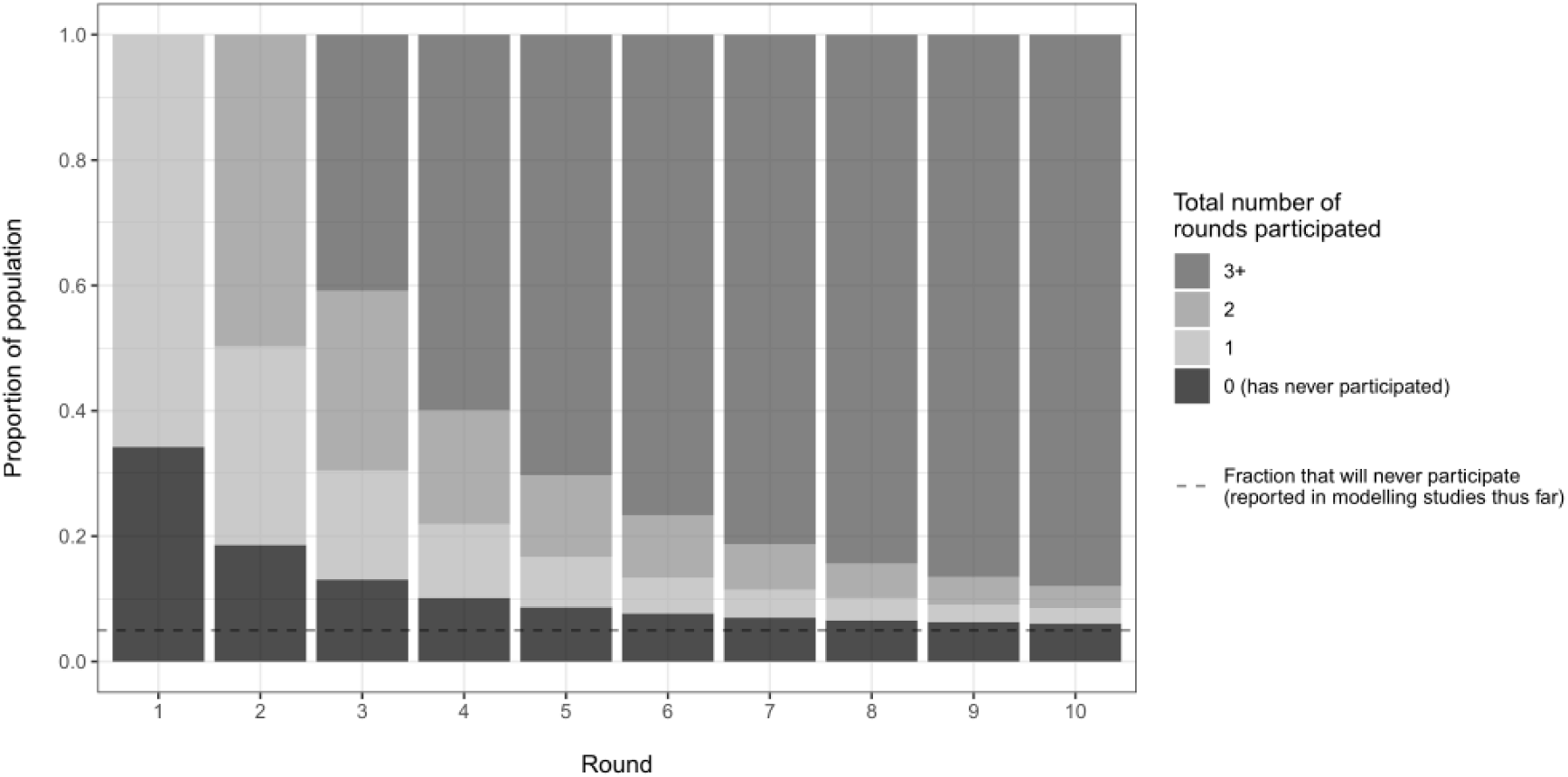
Proportion of the all-time eligible population (y-axis) that has been treated in *N* MDA rounds (grey colour) by the *X*-th round (x-axis) as predicted by the model for MDA participation implemented in LYMFASIM, assuming an annual coverage of 65% of the eligible population, and assuming that 5% will never participate in the long run (dashed horizontal line). The never treated proportion (black) is highest in the first round and declines with the number of treatment rounds, approaching the proportion that will never participate. The all-time eligible population is the subgroup of the population who were eligible for treatment during all *X* treatment rounds.

In the version of TRANSFIL used here, patterns of repeated (non-)treatment are modelled based on Griffin et al. [29] (as reformulated by Dyson et al. [10]). The model contains a parameter, p, which controls the correlation of individuals attending treatment in different rounds. This parameter governs the relative contribution of random and systematic factors to the probability of an individual being treated across consecutive treatment rounds (p = 0 corresponds to completely random and independent probability of attendance in each round; p = 1 corresponds to completely systematic). For TRANSFIL, we simulate different levels of NT by varying the parameter p. In addition to the above, both models consider age-dependent eligibility for treatment.

NT is dynamic over time as it depends on how many rounds have been administered (illustrated in Figure 1). In the current study, we define NT as the proportion of people who are never treated after 5 rounds of MDA among individuals who are eligible (based on their age and health status) for treatment during each of those MDA rounds. We quantify NT after 5 rounds of MDA, as this is the time-point at which most LF programmes evaluate infection prevalence and programme success, and coverage surveys are likely to be implemented. This definition excludes individuals that were or became eligible for treatment during those MDA rounds. The proportion never treated will be higher in young children who were ineligible during all or some of the 5 MDA rounds.

Other than age-dependent eligibility for treatment, no age/sex-specific variation in treatment probabilities (e.g., related to work and mobility or pregnancy status) is considered in the current study. We also assume that ‘drugs received’ are ‘drugs swallowed’ (e.g., there is no coverage-compliance gap). Both models assume no association between treatment and exposure to infection.

### Simulated settings and scenarios

In this analysis we consider two treatment-naïve settings (i.e., without previous history of control by MDA): the first one representing Africa-like populations with *Anopheles gambiae sensu lato* (s.l.)-driven transmission and the second one representing India-like populations with *Culex quinquefasciatus*-driven transmission. The main difference in the models for these two settings is in assumptions around uptake of parasites by the mosquito. In addition, for the Indian setting, the LYMFASIM model considers density-dependent parasite establishment as a result of L3-driven host immunity. For both settings we consider a range of baseline mf prevalence levels in those aged 5 years and older (10%, 20% and 30% in Africa; 5% and 10% in India) and different treatment regimens (IA, DA, or IDA in Africa, and DA, or IDA in India). Table 1 provides an overview of treatment efficacy parameters and age criteria for treatment eligibility considered in the simulations. For all settings, we assume that no bed-nets are implemented. To generate the range of pre-control prevalences, we vary transmission parameters and select simulations and associated parameter values that result in equilibrium prevalences close to the desired baseline mf prevalence (see Supplementary Material for details). The human population size is fixed at 1000.

**Table 1:**
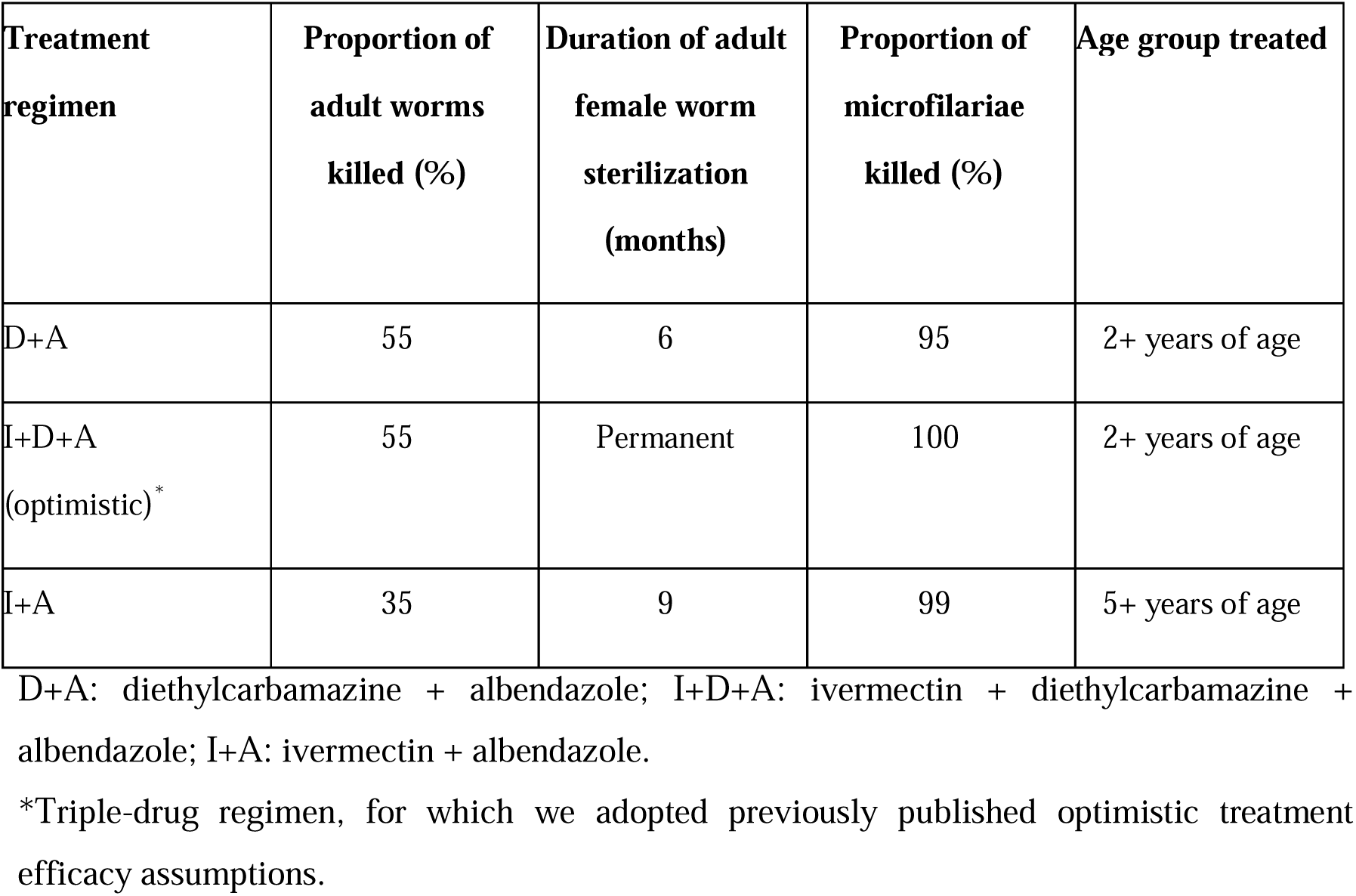
Treatment efficacy assumptions and eligible age groups according to drug combination (taken from [26]).

| **Treatment regimen** | **Proportion of adult worms killed (%)** | **Duration of adult female worm sterilization (months)** | **Proportion of microfilariae killed (%)** | **Age group treated** |
| --- | --- | --- | --- | --- |
| D+A | 55 | 6 | 95 | 2+ years of age |
| I+D+A<br>(optimistic)* | 55 | Permanent | 100 | 2+ years of age |
| I+A | 35 | 9 | 99 | 5+ years of age |

D+A: diethylcarbamazine + albendazole; I+D+A: ivermectin + diethylcarbamazine + albendazole; I+A: ivermectin + albendazole.
\*Triple-drug regimen, for which we adopted previously published optimistic treatment efficacy assumptions.

The impact of NT will depend on its magnitude, as well as the overall population coverage per round and the baseline mf prevalence. Therefore, for an LF program with up to 20 years of annual MDA, we investigated different scenarios with regard to: 1) MDA coverage among the eligible population (Table 1) of 65%, 80%, and 90%. Due to the different age-related eligibility criteria for the drug combinations that can typically be used in each setting, these coverage levels of eligible population correspond, respectively, to 55%-63%, 68%-78%, and 76%-87% among total population for Africa-like or India-like settings. 2) range of NT among eligible individuals varying from 0% to 35% measured after 5 years of MDA.

### Calculation of the probability of mf prevalence **≤** 1%

For all settings, treatment strategies and levels of NT, we calculate the probability of reaching the 1% mf prevalence threshold as the percentage of 500 repeated simulations that achieve the target in the population aged 5 years and above. This percentage is calculated at yearly intervals, just before the next treatment round. We compare the effect of different coverage and NT levels in terms of the number of annual MDA rounds required to achieve the 1% mf prevalence threshold with 90% probability (Figure S1) and identify those scenarios that achieve this within 10 years of annual MDA.

## Results

In Table 2, and Supplementary Material Tables S1⍰S4 we present the number of annual MDA rounds, as a range over both models, required to achieve the 1% mf prevalence threshold with 90% probability for different treatment regimens, MDA coverage, NT, and baseline endemicity levels in treatment-naïve settings. For each setting, we find that: (i) the higher the value of NT (going down the rows in each panel), the higher the number of MDA rounds required for the programme to achieve the target; (ii) the higher the coverage, the fewer the number of rounds required to achieve the target (going across the columns in each panel), and (iii) the higher the baseline mf prevalence (going down the panels), the lower the value of NT tolerable by the programme to achieve the target.

**Table 2:**
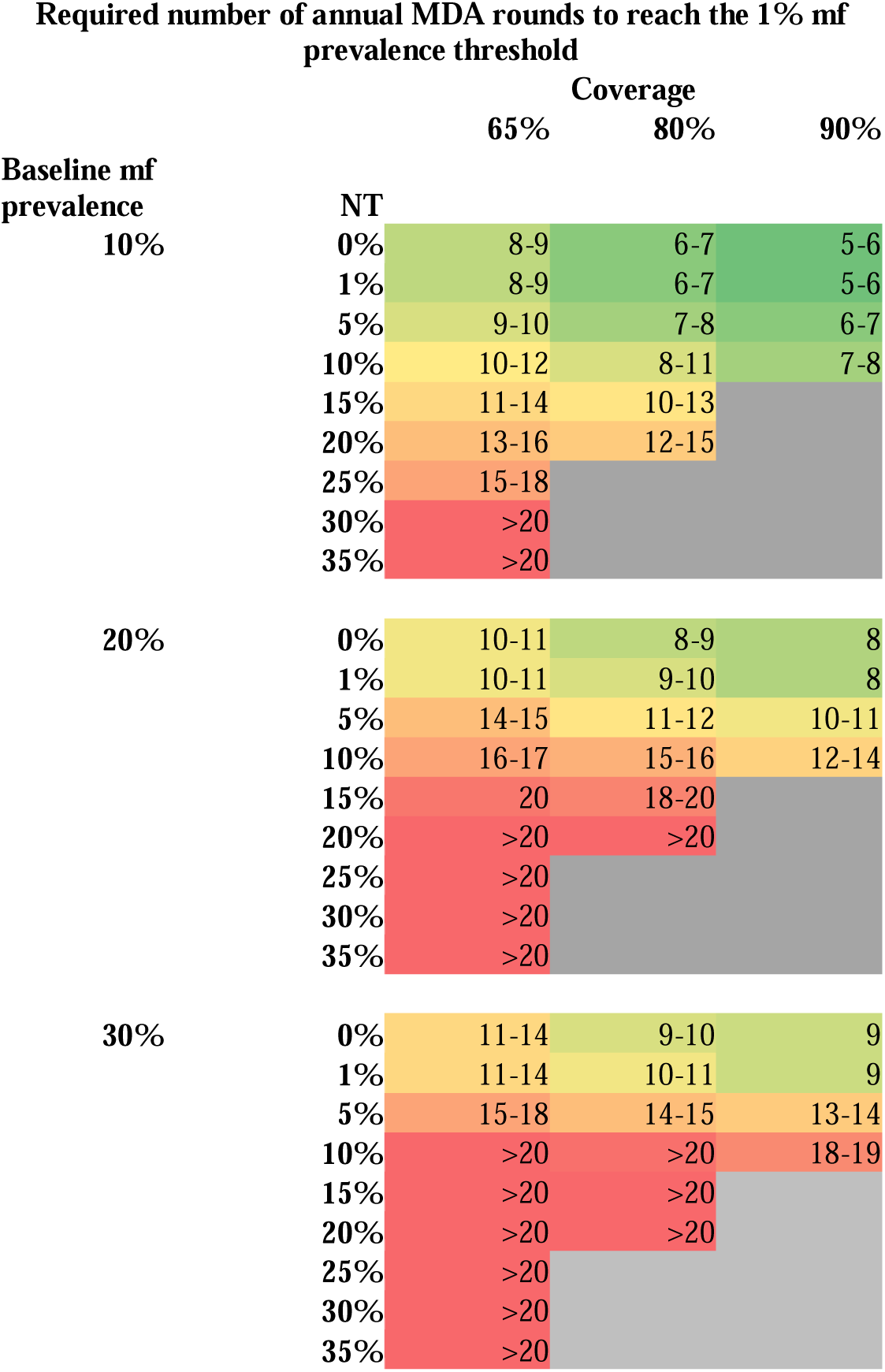
Africa-like settings with anopheline transmission, annual treatment with IA. The number of years, as a range over both models, required to achieve a 90% probability of reaching the 1% mf prevalence threshold interruption of transmission under annual treatment with IA. Coverage and NT levels are among the eligible population. Abbreviations: NT, proportion of the all-time eligible population that has never been treated after 5 rounds of mass drug administration; mf, microfilaremia. Shaded areas = the 1% mf prevalence threshold achieved within 10 years (green), 10-20 years (orange), or >20 years (red); grey shaded areas= scenarios not possible to simulate.

### *Anopheles*-transmission settings

In Africa-like *Anopheles* transmission settings with a baseline mf prevalence of 10% in those aged >5 years and IA treatment at 80% coverage of the eligible population, it is likely that the 1% mf prevalence threshold will be achieved within 10 years of annual treatment when NT <10%. For a baseline mf prevalence between 20% and 30%, this goal could be achieved within a similar timeframe, but only if NT ≤1% (Table 2). This means that in areas with higher pre-control mf prevalence, a nearly perfect MDA coverage would need to be achieved to reach the target within 10 years of MDA treatment. For NT values greater than these levels (10% for a baseline mf prevalence of 10% and 1% for a baseline mf prevalence of 20%⍰30%), 90% probability of <1% mf prevalence can only be reached within 10 years of MDA if treatment coverage is very high (e.g., 90% of the eligible population; Table 2) or if a more effective drug combination were used in areas not co-endemic with onchocerciasis or loiasis (e.g., DA or IDA; Supplementary Material Tables S1⍰S2).

### *Culex*-transmission settings

For India-like settings (with *Culex* vector) with a baseline mf prevalence of 5% in those aged ≥ 5-years, and DA or IDA treatment at 80% coverage of eligibles, it is likely that the 1% mf prevalence threshold will be achieved within 10 years of treatment, regardless of the NT value (up to the value that are feasible to simulate) (Table S3). If coverage level was decreased to 65% of the eligible population, it would be possible to reach <1% mf prevalence within 10 years of annual DA treatment if NT ≤20%. However, treatment with the triple drug (IDA) therapy could achieve this target within 10 years, if NT <25% (Table S3).

In settings with higher mf baseline prevalence (10%), a coverage level of 80% of eligibles is projected to achieve the target in 10-11 years if NT ≤15%, and this outcome applies for both DA and IDA treatment (Table S3 and Table S4). However, the programme duration (number of years required to achieve <1% mf prevalence) is shortest when treating with IDA (Table S3). Treating 90% of the eligible population with DA or IDA can achieve the target within 7 years. By contrast, if coverage were 65%, annual DA treatment could achieve the target within 10 years if NT ≤10%, whilst the tolerable value of NT could increase to ≤15% if IDA were used (Table S3 and Table S4).

## Discussion

Our findings indicate that the level of NT above which the 1% mf prevalence threshold cannot be achieved depends on the baseline endemicity, the employed drug combination, and the MDA coverage levels. The MDA coverage needed to achieve <1% mf prevalence depends on transmission setting, baseline endemicity level, drug combination and the level of NT. In Africa, after five rounds of MDA, the proportion of NT among all-time eligible individuals should not exceed 10% in low endemic settings or 1% in high-endemic settings. The use of more effective drug combinations (DA and IDA compared to IA) can bring these NT thresholds up to some extent, although these cannot be used everywhere (e.g., in areas co-endemic for onchocerciasis or loiasis). In India and other culicine transmission settings, after five rounds of annual MDA at 65% coverage, the proportion of NT should not exceed 20% in low endemic areas. A higher coverage level leads to more optimistic outcomes, in which any NT level can be permitted to achieve <1% mf prevalence. For higher endemic areas, we obtain similar NT thresholds as for the Africa-like settings with anopheline transmission when using the same treatment regimen, coverage level and for a baseline mf prevalence of 10%.

It should be noted that these outcomes are highly dependent on the assumptions that (i) NT occurs completely at random (not clustered in households or sub-communities) and (ii) NT is independent of exposure/infection status. In reality, the first assumption may not necessarily apply: never treated individuals may be clustered geographically, leading to hotspots of ongoing transmission and a larger negative impact of never treatment on required treatment duration.

Regarding the second assumption, the potential (positive or negative) correlation between an individual risk of infection acquisition (e.g., via exposure to mosquito bites) and the probability that an individual has never been treated can influence the impact of NT on achieving the TAS epidemiological thresholds. A positive correlation, where a higher bite-risk corresponds to a higher NT value, decreases the impact of MDA in achieving <1% mf prevalence and would make our results more pessimistic. This is particularly important in settings for which our models predict that the 1% mf prevalence threshold can be achieved with relatively low MDA coverage and high NT values (i.e., in India settings, treating with DA at 65% coverage and 20%NT), which in case of positive correlation, would allow for relatively large reservoirs of infection in untreated people. A negative correlation, where a higher bite-risk corresponds to a low NT, can increase the probability of elimination, and impact the time in achieving the 1% mf prevalence threshold.

Pragmatically, a coverage-NT combination may be acceptable when such a combination permits reaching the 1% mf prevalence threshold, within 10 years of MDA, even though a lower value of NT could have led to achieving the target faster. For example, we can achieve the 1% mf prevalence threshold within 8 years when the annual MDA coverage is 80% and NT is 20% (Table S3-S4). However, we can achieve this target twice as fast when the annual MDA coverage is 80% and NT is 1%. In both scenarios, we use the same number of MDA tablets per round, but over the total programme duration (to reach the 1% mf prevalence threshold) many more tablets are needed when NT is high. Therefore, in order to optimize/prioritize drug use, given limited resources, we need to minimize NT.

Our results can inform policy makers on optimal treatment strategies and show the importance of quantifying the level of NT in a community/implementation unit (and ultimately understanding the reasons behind NT). However, in practice, it might be challenging to quantify NT levels, as it is difficult to identify individuals who have never been treated without implementing longitudinal surveys that record the treatment-related behaviour of each individual during any round of MDA. Although many LF studies have measured the level of NT [30–32], very few longitudinal studies of never treatment have been conducted [33]. Furthermore, most of these studies have measured the level of NT retrospectively and not at each round of MDA, mostly due to financial and logistical constraints. In order to properly model the impact of NT, longitudinal cohort studies with information on who is treated (disaggregated by age, gender, occupation, education) and when, are essential.

It should be noted that there are other factors, not included in this analysis, which can influence the impact of NT on achieving elimination. One of these factors is age/gender-variation in coverage and NT, which depends on age-related patterns of exposure and contribution to transmission and can increase the number of years required to achieve elimination. Here we have explored the impact of MDA as a standalone intervention upon achieving the 1% mf prevalence threshold for a range of NT values. Consideration of other interventions, such as the addition of vector control could alter the results presented here by reducing the vector/human ratio (a component of the vector biting rate, which is a key determinant of the basic reproduction ratio (R_0_) of the infection and hence, of baseline endemicity) and the vector biting rate more generally, helping to decrease the number of years required to achieve the 1% mf prevalence threshold [21].

Human movement (migration) can also play an important role, particularly as elimination is approached, which can either reduce the probability of elimination by adding a source of infective material acting as an external reservoir or increase the probability of elimination by reducing the prevalence of infection in that area [34,35]. This impact depends on human demographic and sociological factors of the area.

## Authors’ contributions

K.K.: conceptualization, formal analysis, investigation, methodology, visualization, project administration, writing—original draft, writing—review and editing; W.A.S.: conceptualization, formal analysis, investigation, methodology, visualization, writing—original draft, writing—review and editing; M.G.B.: conceptualization, methodology, writing—original draft, writing—review and editing; B.S.C.: conceptualization, software, writing—review and editing; S.J.D.V., P.D., K.G.: conceptualization, methodology, writing—review and editing; M.G: conceptualization, software, writing—review and editing; T.D.H., J.D.K., A.K.: conceptualization, methodology, writing—review and editing; R.M.A.: conceptualization, methodology, visualization, supervision, writing—review and editing. L.E.C.: conceptualization, formal analysis, investigation, methodology, visualization, writing—original draft, writing—review and editing.

## Funding

The authors are grateful for funding by the Bill & Melinda Gates Foundation through the NTD Modelling Consortium (grant INV-030046). This supplement is sponsored by funding of Professor T. Deirdre Hollingsworth’s research by Li Ka Shing Foundation at the Big Data Institute, Li Ka Shung Centre for Health Information and Discovery, University of Oxford and funding of the NTD Modelling Consortium by the Bill & Melinda Gates Foundation (INV-030046). K.K., M.G.B., B.S.C., and R.M.A. also acknowledge funding from the MRC Centre for Global Infectious Disease Analysis (MR/R015600/1), jointly funded by the UK Medical Research Council (MRC) and the UK Foreign, Commonwealth & Development Office (FCDO), under the MRC/FCDO Concordat agreement and is also part of the EDCTP2 programme supported by the European Union.

## Data Availability

All data produced in the present work are contained in the manuscript

## Abbreviations

DA: diethylcarbamazine + albendazole
EPHP: elimination as a public health problem
GPELF: Global Programme to Eliminate Lymphatic Filariasis
IA: ivermectin + albendazole
IDA: ivermectin + diethylcarbamazine + albendazole
LF: lymphatic Filariasis
mf: microfilaria
NTD: neglected tropical disease
TAS: transmission assessment survey
WHO: World Health Organization

## Notes

### Competing Interest Statement

The authors have declared no competing interest.

## References

1. World Health Organization. Ending the neglect to attain the Sustainable Development Goals: a road map for neglected tropical diseases 2021–2030. 2021. https://www.who.int/publications/i/item/9789240010352 (accessed 20 June 2023).

2. Zeldenryk LM, Gray M, Speare R, Gordon S, Melrose W. The emerging story of disability associated with lymphatic filariasis: A critical review. PLoS Neglected Tropical Diseases. 2011. doi:10.1371/journal.pntd.0001366

3. Michael E, Bundy DAP, Grenfell BT. Re-assessing the global prevalence and distribution of lymphatic filariasis. Parasitology. 1996. doi:10.1017/s0031182000066646

4. Lenk EJ, Redekop WK, Luyendijk M, Rijnsburger AJ, Severens JL. Productivity Loss Related to Neglected Tropical Diseases Eligible for Preventive Chemotherapy: A Systematic Literature Review. PLoS Neglected Tropical Diseases. 2016. doi:10.1371/journal.pntd.0004397

5. Global programme to eliminate lymphatic filariasis: progress report, 2016. Relev Epidemiol Hebd. 2017.

6. WHO. Global Programme to Eliminate Lymphatic Filariasis: A Manual for National Elimination Programmes. WHO. 2011.

7. Lymphatic filariasis. [cited 24 Jun 2022]. Available: https://www.who.int/news-room/fact-sheets/detail/lymphatic-filariasis. Accessed 1 Dec 2022.

8. Fischer PU, King CL, Jacobson JA, Weil GJ. Potential Value of Triple Drug Therapy with Ivermectin, Diethylcarbamazine, and Albendazole (IDA) to Accelerate Elimination of Lymphatic Filariasis and Onchocerciasis in Africa. PLoS Negl Trop Dis. 2017. doi:10.1371/journal.pntd.0005163

9. World Health Organization. Guideline: Alternative Mass Drug Administration Regimens to Eliminate Lymphatic Filariasis. World Heal Organ. 2017; 1–71. Available: https://www.ncbi.nlm.nih.gov/books/NBK487830/. Accessed 1 Dec 2022.

10. Dyson L, Stolk WA, Farrell SH, Hollingsworth TD. Measuring and modelling the effects of systematic non-adherence to mass drug administration. Epidemics. 2017. doi:10.1016/j.epidem.2017.02.002

11. Krentel A, Gyapong M, Ogundahunsi O, Amuyunzu-Nyamongo M, McFarland DA. Ensuring no one is left behind: Urgent action required to address implementation challenges for NTD control and elimination. PLoS Neglected Tropical Diseases. 2018. doi:10.1371/journal.pntd.0006426

12. King JD, Jacobson J, Krentel A. Accelerating the Uptake of WHO Recommendations for Mass Drug Administration Using Ivermectin, Diethylcarbamazine, and Albendazole. Am J Trop Med Hyg. 2022. doi:10.4269/ajtmh.21-0972

13. Krentel A, Basker N, de Rochars MB, Bogus J, Dilliott D, Direny AN, et al. A multicenter, community-based, mixed methods assessment of the acceptability of a triple drug regimen for elimination of lymphatic filariasis. PLoS Negl Trop Dis. 2021. doi:10.1371/journal.pntd.0009002

14. Krentel A, Fischer PU, Weil GJ. A Review of Factors That Influence Individual Compliance with Mass Drug Administration for Elimination of Lymphatic Filariasis. PLoS Negl Trop Dis. 2013. doi:10.1371/journal.pntd.0002447

15. Understanding who has never been treated during MDA programs – iCHORDS. Available: https://ichords.org/task-teams/understanding-non-adherence-with-mda/. Accessed 1 Dec 2022.

16. Research Links 2 Systematic Non Compliance 13May - YouTube. Available: https://www.youtube.com/watch?v=4wx6sNl6eHs. Accessed 1 Dec 2022.

17. Hollingsworth TD, Adams ER, Anderson RM, Atkins K, Bartsch S, Basáñez MG, et al. Quantitative analyses and modelling to support achievement of the 2020 goals for nine neglected tropical diseases. Parasites and Vectors. 2015. doi:10.1186/s13071-015-1235-1

18. Anderson RM, May RM (Robert M. Infectious diseases of humans[: dynamics and control. Oxford University Press; 1991. Available: https://global.oup.com/academic/product/infectious-diseases-of-humans-9780198540403?cc=gb&lang=en&

19. Irvine MA, Stolk WA, Smith ME, Subramanian S, Singh BK, Weil GJ, et al. Effectiveness of a triple-drug regimen for global elimination of lymphatic filariasis: a modelling study. Lancet Infect Dis. 2017. doi:10.1016/S1473-3099(16)30467-4

20. Irvine MA, Hollingsworth TD. Making Transmission Models Accessible to End-Users: The Example of TRANSFIL. PLoS Negl Trop Dis. 2017. doi:10.1371/journal.pntd.0005206

21. Irvine MA, Reimer LJ, Njenga SM, Gunawardena S, Kelly-Hope L, Bockarie M, et al. Modelling strategies to break transmission of lymphatic filariasis - aggregation, adherence and vector competence greatly alter elimination. Parasit Vectors. 2015. doi:10.1186/s13071-015-1152-3

22. Plaisier AP, Subramanian S, Das PK, Souza W, Lapa T, Furtado AF, et al. The LYMFASIM simulation program for modeling lymphatic filariasis and its control. Methods Inf Med. 1998. doi:10.1055/s-0038-1634505

23. Stolk WA, De Vlas SJ, Borsboom GJJM, Habbema JDF. LYMFASIM, a simulation model for predicting the impact of lymphatic filariasis control: Quantification for African villages. Parasitology. 2008. doi:10.1017/S0031182008000437

24. Subramanian S, Stolk WA, Ramaiah KD, Plaisier AP, Krishnamoorthy K, Van Oortmarssen GJ, et al. The dynamics of Wuchereria bancrofti infection: A model-based analysis of longitudinal data from Pondicherry, India. Parasitology. 2004. doi:10.1017/S0031182004004822

25. Plaisier AP, Stolk WA, Van Oortmarssen GJ, Habbema JDF. Effectiveness of annual ivermectin treatment for Wuchereria bancrofti infection. Parasitology Today. 2000. doi:10.1016/S0169-4758(00)01691-4

26. Stolk WA, Prada JM, Smith ME, Kontoroupis P, De Vos AS, Touloupou P, et al. Are alternative strategies required to accelerate the global elimination of lymphatic filariasis? Insights from mathematical models. Clin Infect Dis. 2018. doi:10.1093/cid/ciy003

27. Farrell SH, Truscott JE, Anderson RM. The importance of patient compliance in repeated rounds of mass drug administration (MDA) for the elimination of intestinal helminth transmission. Parasites and Vectors. 2017. doi:10.1186/s13071-017-2206-5

28. Prada JM, Stolk WA, Davis EL, Touloupou P, Sharma S, Munoz J, et al. Delays in lymphatic filariasis elimination programmes due to COVID-19, and possible mitigation strategies. Trans R Soc Trop Med Hyg. 2021. doi:10.1093/trstmh/trab004

29. Griffin JT, Hollingsworth TD, Okell LC, Churcher TS, White M, Hinsley W, et al. Reducing Plasmodium falciparum malaria transmission in Africa: A model-based evaluation of intervention strategies. PLoS Med. 2010. doi:10.1371/journal.pmed.1000324

30. Niles RA, Thickstun CR, Cox H, Dilliott D, Burgert-Brucker CR, Harding-Esch EM, et al. Assessing factors influencing communities’ acceptability of mass drug administration for the elimination of lymphatic filariasis in guyana. PLoS Negl Trop Dis. 2021. doi:10.1371/journal.pntd.0009596

31. Krentel A, Damayanti R, Titaley CR, Suharno N, Bradley M, Lynam T. Improving Coverage and Compliance in Mass Drug Administration for the Elimination of LF in Two ‘Endgame’ Districts in Indonesia Using Micronarrative Surveys. PLoS Negl Trop Dis. 2016. doi:10.1371/journal.pntd.0005027

32. El-Setouhy M, Abd Elaziz KM, Helmy H, Farid HA, Kamal HA, Ramzy RMR, et al. The effect of compliance on the impact of mass drug administration for elimination of lymphatic filariasis in Egypt. Am J Trop Med Hyg. 2007. doi:10.4269/ajtmh.2007.77.1069

33. Shuford K V., Turner HC, Anderson RM. Compliance with anthelmintic treatment in the neglected tropical diseases control programmes: A systematic review. Parasites and Vectors. 2016. doi:10.1186/s13071-016-1311-1

34. Ramaiah KD. Population migration: implications for lymphatic filariasis elimination programmes. PLoS Negl Trop Dis. 2013;7. doi:10.1371/JOURNAL.PNTD.0002079

35. Vegvari C, Truscott JE, Kura K, Anderson RM. Human population movement can impede the elimination of soil-transmitted helminth transmission in regions with heterogeneity in mass drug administration coverage and transmission potential between villages: A metapopulation analysis. Parasites and Vectors. 2019. doi:10.1186/s13071-019-3612-7

