## Supplemental file for "How does the proportion of never treatment influence the success of mass drug administration programmes for the elimination of lymphatic filariasis?"

### 1. Additional results

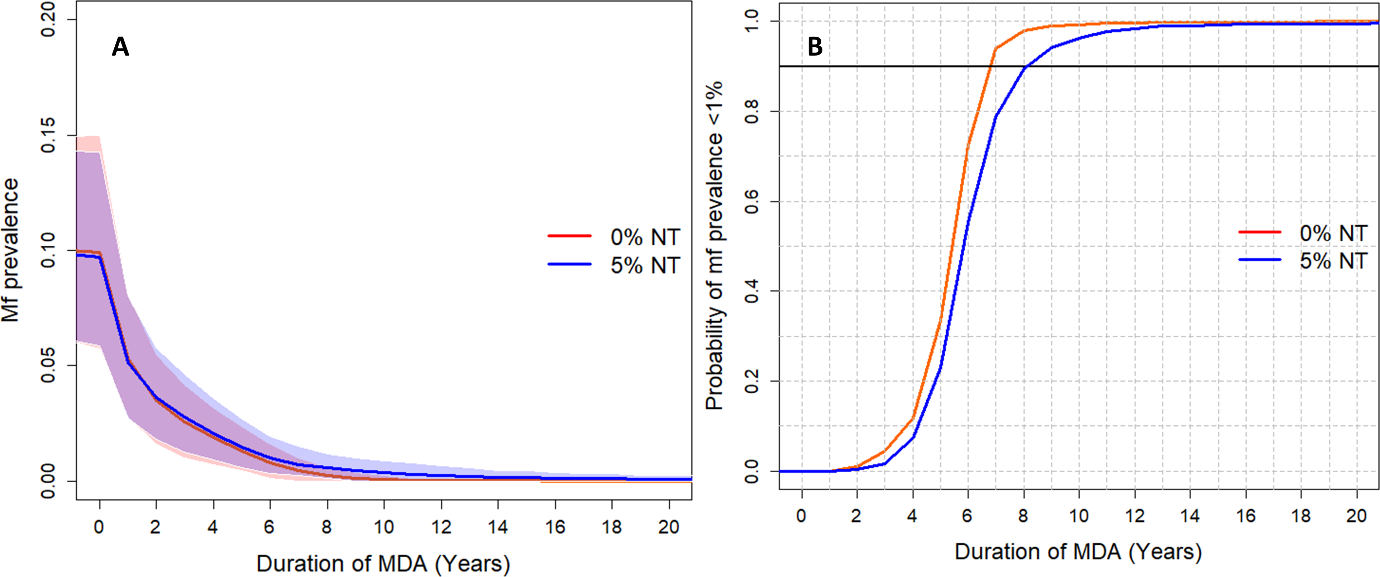

**Figure S1:** Example of Wuchereria bancrofti microfilaraemia (Mf) prevalence trends (in those aged ≥5 years) over 20 years of treatment (A) and the probability of achieving the 1% mf prevalence threshold (B) for Africa-like settings with Anopheles transmission, when the baseline prevalence is 10% and 80% of the eligible population is annually treated with ivermectin+albendazole (IA). The red line corresponds to 0% proportion of individuals never treated (NT) (i.e., no individuals are permanently excluded from treatment). The blue line indicates 5% NT. Shaded areas (both red and blue, respectively) represent the 90% prediction interval credible interval (90% of the simulated results fall within these shaded areas). The horizontal black line in (B) indicates 90% probability of reaching the target. Results are generated using the TRANSFIL model.

**Table S1: Africa-like settings with anopheline transmission, annual treatment with DA.** The number of years, as a range over both models, required to achieve a 90% probability of reaching EPHP under annual treatment with DA. Coverage and NT levels are among the eligible population. Abbreviations: NT, proportion of the all-time eligible population that has never been treated after 5 rounds of mass drug administration; EPHP, elimination as a public health problem (considered achieved if the mf prevalence reaches <1% mf prevalence); mf, microfilarial. Shaded areas = EPHP achieved within 10 years (green), 10-20 years (yellow-orange), or >20 years (red); grey shaded areas= scenarios not possible.

| **Required treatment duration (in years) to reach EPHP (<1% mf prevalence)** | | | | | | |
| --- | --- | --- | --- | --- | --- | --- |
|  |  | **Coverage** | | | | |
|  |  | **65%** | | **80%** | | **90%** |
| **Baseline mf prevalence** | **NT** |  | |  | |  |
| **10%** | **0%** | 6-7 | | 5-6 | | 4-5 |
|  | **1%** | 6-8 | | 5-6 | | 4-5 |
|  | **5%** | 7-9 | | 6-7 | | 5-6 |
|  | **10%** | 9-12 | | 7-9 | | 6-7 |
|  | **15%** | 10-14 | | 10-11 | |  |
|  | **20%** | 12-18 | | 12-13 | |  |
|  | **25%** | 15-20 | |  | |  |
|  | **30%** | 16-20 | |  | |  |
|  | **35%** | >20 | |  | |  |
| **20%** | **0%** | 8-9 | | 6-7 | | 6 |
|  | **1%** | 8-9 | | 7 | | 6 |
|  | **5%** | 12 | | 10 | | 8-9 |
|  | **10%** | 15-16 | | 14-15 | | 11-14 |
|  | **15%** | 18-20 | | 17-18 | |  |
|  | **20%** | >20 | | >20 | |  |
|  | **25%** | >20 | |  | |  |
|  | **30%** | >20 | |  | |  |
|  | **35%** | >20 | |  | |  |
| **30%** | **0%** | 9-10 | | 7-8 | | 6 |
|  | **1%** | 9-10 | | 8 | | 7 |
|  | **5%** | 14-15 | | 12-13 | | 10-13 |
|  | **10%** | 18-20 | | 16-18 | | 14-18 |
|  | **15%** | >20 | | 20 | |  |
|  | **20%** | >20 | | >20 | |  |
|  | **25%** | >20 | |  | |  |
|  | **30%** | >20 | |  | |  |
|  | **35%** | >20 | |  | |  |

**Table S2: Africa-like settings with anopheline transmission, annual treatment with IDA.** The number of years, as a range over both models, required to achieve a 90% probability of reaching EPHP under annual treatment with IDA. Coverage and NT levels are among the eligible population. Abbreviations: NT, proportion of the all-time eligible population that has never been treated after 5 rounds of mass drug administration; EPHP, elimination as a public health problem; mf, microfilarial. Shaded areas = EPHP achieved within 10 years (green), 10-20 years (yellow-orange), or >20 years (red); grey shaded areas= scenarios not possible.

| **Required treatment duration (in years) to reach EPHP (<1% mf prevalence)** | | | | |
| --- | --- | --- | --- | --- |
|  |  | **Coverage** | | |
|  |  | **65%** | **80%** | **90%** |
| **Baseline mf prevalence** | **NT** |  |  |  |
| **10%** | **0%** | 4-6 | 2-5 | 2-4 |
|  | **1%** | 4-6 | 2-5 | 2-4 |
|  | **5%** | 6-7 | 4-6 | 2-5 |
|  | **10%** | 9 | 6-7 | 3-7 |
|  | **15%** | 10-11 | 8-10 |  |
|  | **20%** | 12-15 | 10-12 |  |
|  | **25%** | 15-18 |  |  |
|  | **30%** | 16-20 |  |  |
|  | **35%** | >20 |  |  |
| **20%** | **0%** | 6-8 | 3-6 | 2-6 |
|  | **1%** | 6-8 | 4-7 | 2-6 |
|  | **5%** | 9-12 | 7-10 | 5-9 |
|  | **10%** | 13-15 | 11-15 | 8-14 |
|  | **15%** | 18 | 14-18 |  |
|  | **20%** | 20 | 20 |  |
|  | **25%** | >20 |  |  |
|  | **30%** | >20 |  |  |
|  | **35%** | >20 |  |  |
| **30%** | **0%** | 7-10 | 5-8 | 2-6 |
|  | **1%** | 7-10 | 5-8 | 3-7 |
|  | **5%** | 10-15 | 9-13 | 8-13 |
|  | **10%** | 15-20 | 14-18 | 11-18 |
|  | **15%** | >20 | 18-20 |  |
|  | **20%** | >20 | >20 |  |
|  | **25%** | >20 |  |  |
|  | **30%** | >20 |  |  |
|  | **35%** | >20 |  |  |

**Table S3: India-like settings with culicine transmission, annual treatment with IDA.** The number of years, as a range over both models, required to achieve a 90% probability of reaching EPHP under annual treatment with IDA. Coverage and NT levels are among the eligible population. Abbreviations: NT, proportion of the all-time eligible population that has never been treated after 5 rounds of mass drug administration; EPHP, elimination as a public health problem; mf, microfilarial. Shaded areas = EPHP achieved within 10 years (green), 10-20 years (yellow-orange), or >20 years (red); grey shaded areas= scenarios not possible.

| **Required treatment duration in years to reach EPHP (<1% mf prevalence)** | | | | |
| --- | --- | --- | --- | --- |
|  |  | **Coverage** | | |
|  |  | **65%** | **80%** | **90%** |
| **Baseline mf prevalence** | **NT** |  |  |  |
| **5%** | **0%** | 2-3 | 2 | 1 |
|  | **1%** | 2-3 | 2 | 1 |
|  | **5%** | 3 | 2 | 1 |
|  | **10%** | 4 | 2-3 | 1 |
|  | **15%** | 6 | 3-5 |  |
|  | **20%** | 8 | 4-8 |  |
|  | **25%** | 10-11 |  |  |
|  | **30%** | 12-18 |  |  |
|  | **35%** | 12-20 |  |  |
| **10%** | **0%** | 3 | 2 | 2 |
|  | **1%** | 3 | 2-3 | 2 |
|  | **5%** | 5 | 3 | 2 |
|  | **10%** | 7 | 6-7 | 4-7 |
|  | **15%** | 9-10 | 8-10 |  |
|  | **20%** | 12-13 | 9-12 |  |
|  | **25%** | 15-17 |  |  |
|  | **30%** | 18-20 |  |  |
|  | **35%** | 18-20 |  |  |

**Table S4: India-like settings with culicine transmission, annual treatment with DA.** The number of years, as a range over both models, required to achieve a 90% probability of reaching EPHP under annual treatment with DA. Coverage and NT levels are among the eligible population. Abbreviations: NT, proportion of the all-time eligible population that has never been treated after 5 rounds of mass drug administration; mf, microfilaremia. Shaded areas = the 1% mf prevalence threshold achieved within 10 years (green), 10-20 years (orange), or >20 years (red); grey shaded areas= scenarios not possible to simulate.

| **Required number of annual MDA rounds to reach the 1% mf prevalence threshold** | | | | |
| --- | --- | --- | --- | --- |
|  |  | **Coverage** | | |
|  |  | **65%** | **80%** | **90%** |
| **Baseline mf prevalence** | **NT** |  |  |  |
| **5%** | **0%** | 4-5 | 3-4 | 3-4 |
|  | **1%** | 4-5 | 3-4 | 3-4 |
|  | **5%** | 5-6 | 4-5 | 3-4 |
|  | **10%** | 6-7 | 4-5 | 3-4 |
|  | **15%** | 7-8 | 6 |  |
|  | **20%** | 9-10 | 6-8 |  |
|  | **25%** | 11-12 |  |  |
|  | **30%** | 13-17 |  |  |
|  | **35%** | 13-20 |  |  |
| **10%** | **0%** | 5-6 | 4-5 | 3-4 |
|  | **1%** | 5-6 | 4-5 | 4 |
|  | **5%** | 7 | 5-6 | 4-5 |
|  | **10%** | 9 | 7-8 | 6-7 |
|  | **15%** | 11 | 10-11 |  |
|  | **20%** | 14 | 11-13 |  |
|  | **25%** | 17-19 |  |  |
|  | **30%** | >20 |  |  |
|  | **35%** | >20 |  |  |

### 2. TRANSFIL model description and methods

### 2.1. Description of the mathematical model

The mathematical model of lymphatic filariasis (LF) transmission TRANSFIL is a stochastic individual-based model of LF infection in human populations. A full model description is given in Irvine et al [1] and more recently in Michael et al [2], so here we provide a brief summary of the model development. TRANSFIL is a stochastic individual-based model, simulating worm burden, microfilaraemia and other demographic parameters relating to age and risk of exposure. Humans are modelled individually, with their own male and female worm burden. The concentration of mf in the peripheral blood is modelled for each individual and increases according to the number of fertile female worms as well as decreasing at a constant rate. The total mf density in the population contributes towards the current density of L3 larvae in the human-biting mosquito population, where the distribution of L3 amongst the human-biting mosquito population is completely homogeneous. An empirically derived relationship is used for the uptake of mf by a mosquito, where both Culex and Anopheles uptake curves are implemented depending on setting (see Irvine et al [1]). The model dynamics are therefore divided into the individual human dynamics, including age and turnover; worm dynamics inside the host; microfilariae dynamics inside the host and larvae dynamics inside the mosquito.

### 2.2. Model implementation

We performed simulations for LF endemic settings in India and Africa, considering a range of precontrol mf prevalence levels (prevalence range 0 to 40% in the scenarios considered) and regional differences in local vector species and standard treatment regimens. More specifically, in order to generate the required range of mf prevalences in individuals above 5 years of age, we varied three parameters of the model, the vector to host ratio (V/H), the average population bite risk (k) and the importation rate (Imp), using parameter sets from a range of plausible values based on previously analysed data [1,2,3]. V/H is an alternative metric to the basic reproductive number ($R_{0}$), which can range from zero to 2.5 [16]. The graphical representation of the values is shown in Figures S1-S3. For stochastic models it is essential that an importation rate is included, otherwise the equilibrium distribution (steady state) that is used as the starting point of the simulations can potentially converge to the degenerate distribution where no-one is infected. The importation rate does not need to be large, in fact it should not be driving the infection. For this LF study we used a random number drawn from a uniform distribution with minimum 0 and maximum 0.00025 (max 2.5/10000 infections per month). The interventions reduce the prevalence over time, and therefore as years pass, the importation rate decreases in proportion to the reduction in prevalence seen in some pilot simulations. Finally, compliance between rounds of MDA is modelled based on the paper by Griffin et al. [14], following the description in Dyson et al. [15] and previous implementation of the model [2]. A summary of all model parameters is available in Table S5.

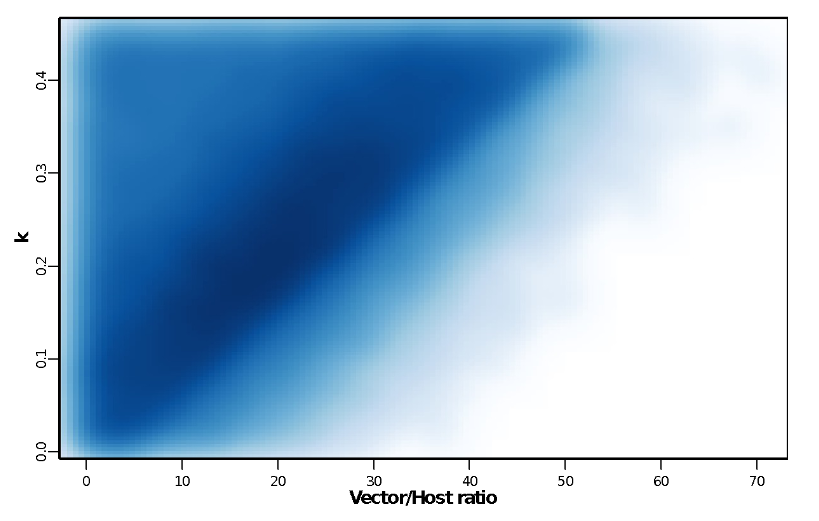

**Figure S2**: Vector to Host ratio against aggregation parameter k. The density plot indicates the parameter space areas from the simulations that were in the 0 to 40% mf prevalence range at baseline. Dark blue areas denote more common parameter values.

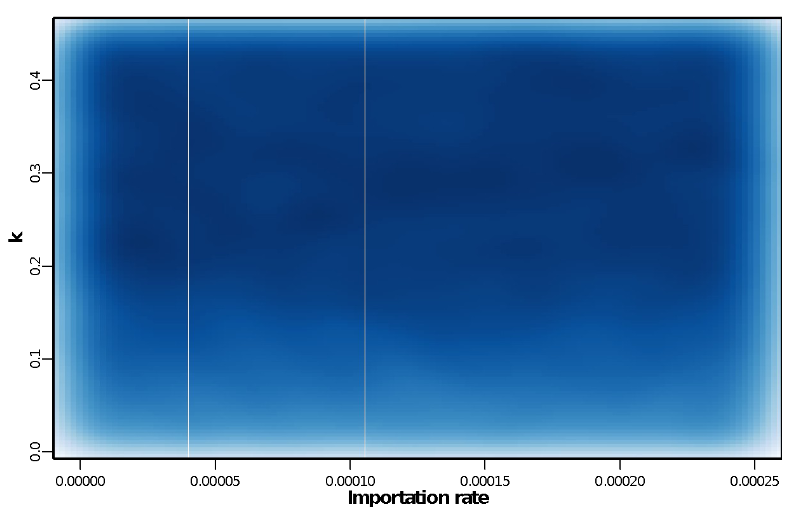

**Figure S3**: Importation rate against aggregation parameter k. The density plot indicates the parameter space areas from the simulations that were in the 0 to 40% mf prevalence range at baseline. Dark blue areas denote more common parameter values.

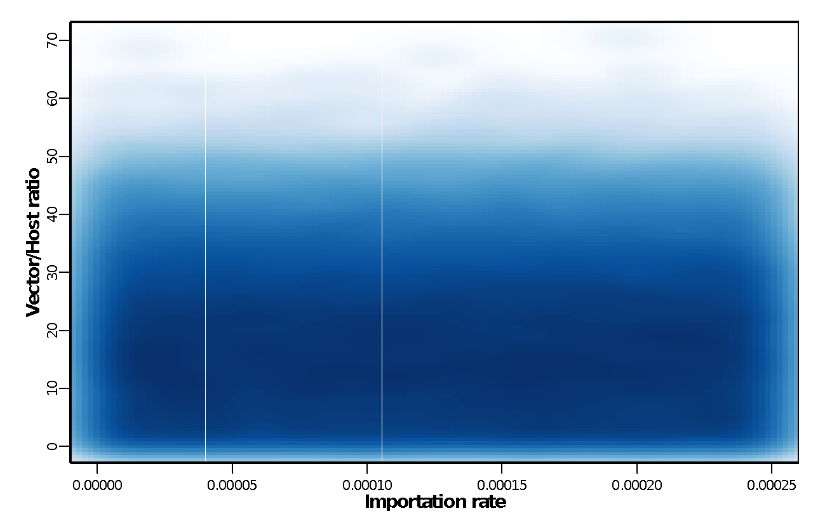

**Figure S4**: Importation rate against Vector to Host ratio. The density plot indicates the parameter space areas from the simulations that were in the 0 to 40% mf prevalence range at baseline. Dark blue areas denote more common parameter values.

**Table S5**: Description of the basic LF model parameters.

| **Parameter symbol** | **Definition** | **Value** | **Source** |
| --- | --- | --- | --- |
| $\lambda$ | Number of bites per mosquito | 10 per month | [4,5] |
| $V/H$ | Ratio of number of vectors to hosts | Varied | Input |
| $\alpha_{max}$ | Age at which exposure to mosquitoes reaches its maximum level | 20 | [6] |
| $\psi_{1}$ | Proportion of L3 leaving mosquito per bite | 0.414 | [7] |
| $\psi_{2}$ | Proportion of L3 leaving mosquito that enter host | 0.32 | [8] |
| $s_{2}$ | Proportion of L3 entering host that develop into adult worms | 0.00275 | [9,10] |
| $\mu$ | Death rate of adult worms | 0.0104 per month | [11] |
| $\delta$ | Production rate of mf per worm | 0.2 per month | [7] |
| $\zeta$ | Death rate f mf | 0.1 per month | [7,12] |
| $g$ | Proportion of mosquitoes which pick up infection when biting an infected host | 0.37 | [13] |
| $\sigma$ | Death rate of mosquitoes | 5 per month | [8] |
| $k$ | Aggregation parameter of individual exposure to mosquitoes | Varied | Input |
| $h(\alpha)$ | Parameter to adjust rate at which individuals of age α are bitten | Linear from 0 to 10, with maximum of 1 | [9] |
| $Imp$ | Importation rate |  | Input |

### 2.3. Modelling never treatment

The model contains a parameter, $\rho$, which controls the correlation of individuals attending treatment in different rounds. This parameter governs the relative contribution of random and systematic factors to the probability of an individual being treated across consecutive treatment rounds ($\rho=0$ corresponds to completely random and independent probability of attendance in each round, $\rho=1$ corresponds to completely systematic). For TRANSFIL, we simulate different levels of NT by varying the parameter $\rho$ (Table S6).

**Table S6**: Parametrization of never treatment (TRANSFIL model) after five rounds of treatment.

| **NT** | **Coverage** | | |
| --- | --- | --- | --- |
|  | **65%** | **80%** | **90%** |
|  | $\boldsymbol{\rho}$ | $\boldsymbol{\rho}$ | $\boldsymbol{\rho}$ |
| 0.01 | 0.07 | 0.31 | 0.59 |
| 0.02 | 0.16 | 0.47 | 0.73 |
| 0.03 | 0.24 | 0.57 | 0.81 |
| 0.04 | 0.31 | 0.64 | 0.87 |
| 0.05 | 0.37 | 0.69 | 0.92 |
| 0.06 | 0.43 | 0.73 | 0.95 |
| 0.07 | 0.47 | 0.77 | 0.97 |
| 0.08 | 0.52 | 0.81 | 0.99 |
| 0.09 | 0.56 | 0.85 | 1 |
| 0.1 | 0.6 | 0.89 | 1 |
| 0.11 | 0.63 | 0.92 |  |
| 0.12 | 0.66 | 0.95 |  |
| 0.13 | 0.69 | 0.97 |  |
| 0.14 | 0.72 | 0.97 |  |
| 0.15 | 0.75 | 0.97 |  |
| 0.16 | 0.78 | 0.96 |  |
| 0.17 | 0.8 | 0.94 |  |
| 0.18 | 0.83 | 0.95 |  |
| 0.19 | 0.85 | 0.98 |  |
| 0.2 | 0.87 | 1 |  |
| 0.21 | 0.89 |  |  |
| 0.22 | 0.91 |  |  |
| 0.23 | 0.92 |  |  |
| 0.24 | 0.93 |  |  |
| 0.25 | 0.95 |  |  |
| 0.26 | 0.96 |  |  |
| 0.27 | 0.96 |  |  |
| 0.28 | 0.97 |  |  |
| 0.29 | 0.97 |  |  |
| 0.3 | 0.98 |  |  |
| 0.31 | 0.98 |  |  |
| 0.32 | 0.99 |  |  |
| 0.33 | 0.99 |  |  |
| 0.34 | 1 |  |  |
| 0.35 | 1 |  |  |

### 2.4.  Model code

The TRANSFIL model code has been made available by the NTD Modelling Consortium

(<https://github.com/sempwn/LF-model>).

### 2.5. References

1. Irvine MA, Reimer LJ, Njenga SM, Gunawardena S, Kelly-Hope L, Bockarie M, and Hollingsworth TD (2015) Modelling strategies to break transmission of lymphatic filariasis - aggregation, adherence and vector competence greatly alter elimination. Parasites and Vectors 8:547.

2. Michael E, Sharma S, Smith ME, Touloupou P, Giardina F, Prada JM, Stolk WA, Hollingsworth TD, de Vlas SJ (2018) Quantifying the value of surveillance data for improving model predictions of lymphatic filariasis elimination. PLoS Negl Trop Dis 12(10): e0006674.

3. Irvine, M.A., Stolk, W.A., Smith, M.E., Subramanian, S., Singh, B.K., Weil, G.J., Michael, E., Hollingsworth, T.D. (2017) Effectiveness of a triple-drug regimen for global elimination of lymphatic filariasis: a modelling study. Lancet Infectious Diseases 17(4), 451–458.

4. Rajagopalan P (1980) Population dynamics of culex pipiens fatigans, the filariasis vector, in pondicherry: influence of climate and environment. Proc Indian Natl Sci Acad. 6: 745–752.

5. Subramanian S, Manoharan A, Ramaiah K, Das P (1994) Rates of acquisition and loss of wuchereria bancrofti infection in culex quinquefasciatus. Am J Trop Med Hyg. 51: 244–249.

6. Subramanian S, Stolk W, Ramaiah K, Plaisier A, Krishnamoorthy K, Van Oortmarssen G (2004). The dynamics of wuchereria bancrofti infection: a model-based analysis of longitudinal data from Pondicherry, India. Parasitology 128: 467–482.

7. Hairston NG, de Meillon B (1968) On the inefficiency of transmission of wuchereria bancrofti from mosquito to human host. Bull World Health Organ. 38: 935.

8. Ho BC, Ewert A (1967) Experimental transmission of filarial larvae in relation to feeding behaviour of the mosquito vectors. Trans R Soc Trop Med Hyg. 61: 663–666.

9. Norman R, Chan MS, Srividya A, Pani S, Ramaiah KD, Vanamail P (2000) EPIFIL: The development of an age-structured model for describing the transmission dynamics and control of lymphatic filariasis. Epidemiol Infect. 124: 529–541.

10. Stolk WA, De Vlas SJ, Borsboom GJ, Habbema J (2008) LYMFASIM, a simulation model for predicting the impact of lymphatic filariasis control: Quantification for African villages. Parasitology 135L: 1583–1598.

11. Evans DB, Gelband H, Vlassoff C (1993) Social and economic factors and the control of lymphatic filariasis: a review. Acta Trop. 53: 1–26.

12. Ottesen E, Ramachandran C (1995) Lymphatic filariasis infection and disease: control strategies. Parasitol Today 11: 129–130.

13. Subramanian S, Krishnamoorthy K, Ramaiah K, Habbema J, Das P, Plaisier A (1998) The relationship between microfilarial load in the human host and uptake and development of wuchereria bancrofti microfilariae by culex quinquefasciatus: a study under natural conditions. Parasitology. 116: 243–255.

14. Griffin JT, Hollingsworth TD, Okell LC, Churcher TS, White M, Hinsley W, Bousema T, Drakeley CJ, Ferguson NM, Basez MG, and Ghani AC. (2010) Reducing Plasmodium falciparum malaria transmission in Africa: a model-based evaluation of intervention strategies. PLOS Medicine. 7: 1-17.

15. Dyson L, Stolk WA, Farrell SH, and Hollingsworth TD (2017) Measuring and modelling the effects of systematic non-adherence to mass drug administration. Epidemics 18: 56-66.

16. Stone C. et al (2014) How effective is integrated vector management against malaria and lymphatic filariasis where the diseases are transmitted by the same vector? PLoS Negl. Trop. Dis. 8, e3393

### 3. LYMFASIM model description and methods

### 3.1. Description of the mathematical model

The LYMFASIM model has been described elsewhere [1,3] and it has been applied to support decision making on control and elimination of lymphatic filariasis in different settings [3-6]. We restrict here to a brief description.

LYMFASIM is a stochastic individual-based model for simulating lymphatic filariasis (LF) transmission and control in a closed, dynamic population, typically representing the population from a village or small town. Each human individual is simulated separately. The population composition changes over time, because of birth, death and emigration (removal) of individuals from the population. The infection status (number of adult worms for each sex, mf density) for each individual in the population is tracked over time. Exposure to mosquito bites is assumed to vary between individuals, driven by age and sex patterns in exposure as well as by stochastic variation between individuals. As a result, infection levels vary between individuals. Female adult worms produce microfilariae (mf) when at least one male worms is present in the same host (polygamous mating). The uptake and transmission of infection between hosts are simulated deterministically, accounting for the variation in exposure between individuals.

LYMFASIM can be used to simulate the effect of interventions (e.g. mass drug administration, integrated vector management, bednet use) on transmission and morbidity, taking account of the human demography and the complexities of helminth transmission. Mass drug administration (MDA) is simulated by specifying the year and month in which treatment takes place, the efficacy of the applied treatment regimen, the achieved coverage level, and compliance patterns. Systematic non-participation is simulated by assuming that a fraction of the population never participates in MDA (e.g. systematic refusal, related to chronic illness). In addition, LYMFASIM allows the relative compliance to vary between age and sex groups; this mechanism captures transient contra-indications for MDA (e.g. exclusion of young children and pregnant women) and other age- and sex-related behavioural factors driving participation in MDA. Lastly, each individual has a personal inclination to participate in MDA, which is considered as a lifelong property. A stochastic process eventually defines per individual whether he is treated in a given round, depending on the calculated probability.

### 3.2. Model parameters and their values

We previously derived model quantifications for simulating transmission of bancroftian filariasis by *Culex* species in India [3] and *Anopheles* species in Africa[7], accounting for the age-structure of the human population and density dependence in the L3 yield from a blood meal in mosquitoes. Acquired immunity was not considered to play a role in the Africa model [7]. Values of parameters that were not varied between simulations are listed in Table S7 below.

Assumptions and parameters related to the treatment efficacy of different employed treatment regimens are listed in the main text. We assume that the mf production resumes immediately after the end of the period of recovery, by assuming a very high value (1000) for the shape parameter of the recovery function.

**Table S7:** LYMFASIM input: probability distributions, functions and parameter values

| **Parameter description (symbol)** | **Model variant for Africa** | **Source / remarks** | **Model variant for India** | **Source / remarks** |
| --- | --- | --- | --- | --- |
| **Human demography** |  |  |  |  |
| Cumulative survival (F(a)), by age (no difference assumed between sexes) | Age Survival  0 1  5 0.800  15 0.79  20 0.755  25 0.737  30 0.723  35 0.654  40 0.605  45 0.560  50 0.506  60 0.487  70 0.305  80 0.155  99 0 | [7], modified as in [8] | Age Survival  0 1  5 0.904  10 0.895  15 0.888  20 0.879  25 0.864  30 0.849  40 0.812  50 0.756  90 0 | [3] |
| Fertility rate per woman (R(a)), by age | Age Fertility rate  0 0  5 0  15 0  20 0.116  25 0.230  30 0.245  35 0.207  40 0.147  45 0.077  50 0.031  60 0  70 0  80 0  99 0 | [7] | Age Fertility rate  0 0  5 0  10 0  15 0  20 0.075  25 0.254  30 0.222  40 0.096  50 0.013  90 0 | [3] |
| Initial population | Age Males/females  5 42/42  15 63/63  20 26/26  25 22/22  30 20/20  35 17/17  40 14/14  45 11/11  50 9/9  60 14/14  70 9/9  80 3/3  99 1/1 | Assumed | Age Males/females  5 20/20  10 17/17  15 15/15  20 15/15  25 22/22  30 20/20  40 15/15  50 13/13  90 13/13 | Assumed |
| Maximum population size | Fixed at 1025 | Assumed | Fixed at 1025 | Assumed |
| Proportion removed when maximum population size is reached | 5% | Assumed | 5% | Assumed |
| **Morbidity and excess mortality from mortality** | Not considered in this simulation | | Not considered in this simulation | |
| **Transmission initialization** |  |  |  |  |
| External force-of-infection at start of burn-in period | 2 | Assumed | 2 | Assumed |
| Duration of external force-of-infection at start of burn-in period | 2 years | Assumed | 2 years | Assumed |
| Duration of warming up period | 159 | Assumed | 159 | Assumed |
| **Transmission dynamics after initalization** |  |  |  |  |
| Average mosquito biting rate for adult men (mbr) | Varied between simulations as described below |  | Varied between simulations as described below |  |
| Seasonal variation | No seasonal variation (monthly biting rate is the same in all months) | Assumed | No seasonal variation (monthly biting rate is the same in all months) | Assumed |
| Relative biting rate (multiplier of mbr that can be used scale a seasonal pattern to some desired level) | 1 |  | 1 |  |
| Variation in exposure by age (no difference assumed between sexes) | 0 at birth, linearly increasing to reach 1 at the age of 20 years and constant at 1 from this age onwards | Previously estimated by fitting to data[3]; slightly adjusted for Africa [7] | 0.26 at birth, linearly increasing to reach 1 at the age of 19.1 years and constant at 1 from this age onwards | Previously estimated by fitting the model to data [3] |
| Probability distribution describing variation in the individual exposure index (Ei), due to personal factors (fixed through life) given age and sex | Gamma distribution with mean 1.0; shape (=rate) is varied as described below | Assumed | Gamma distribution with mean 1.0; shape (=rate) is varied as described below | Assumed |
| External force of infection | 0 |  |  |  |
| **Parasite dynamics within host** |  |  |  |  |
| Success ratio (sr) | 0.00088 | Previously estimated by fitting to data [7] | 0.00103 | Previously estimated by fitting the model to data [3] |
| Anti-L3 immunity | Not considered in this model variant (strength of immunological response = 0; duration of immunological memory for anti-L3 immunity = 0) | Assumed, as justified in [7] | Included in model (strength of immunological response = 0.0000589; duration of immunological memory=9.6 years; Shape-parameter for the gamma-distribution describing  individual variation in the ability to develop an anti-L3  immune-response = 1.07) | Previously estimated by fitting the model to data [3] |
| Anti-fecundity immunity: | Not considered in this model variant (strength of immunological memory for anti-fecundity immunity = 0; duration of immunological memory for anti-fecundity immunity = 0) | Assumed, as justified in [7] | Not considered in this model variant (strength of immunological memory for anti-fecundity immunity = 0; duration of immunological memory for anti-fecundity immunity = 0) |  |
| Average worm lifespan (Tl) | 10 years on average; varied according to a Weibull distribution with shape 2 | Previously estimated by fitting to data [3] | 10.2 years on average; varied according to a Weibull distribution with shape 2 | [3] |
| Duration of immature stage of the parasite in human host (Ti) | Constant, 8 months | Fixed [9] | Constant, 8 months | Fixed [9] |
| No. of Mf produced per female parasite per month per 20 ml peripheral blood in the absence of immune reactions and in the presence of at least 1 male worm (r0) | 0.58 | Previously estimated by fitting to data [7] | 0.606 | Previously estimated by fitting to data [3] |
| Monthly survival of the microfilariae, fraction (s) | 0.9 | Fixed, based on [10] | 0.9 | Fixed, based on [10] |
| Association between worm age and mf production rate | Mf production independent of worm age | Assumed | Mf production independent of worm age | Assumed |
| Polygamy (all female worms produce mf in the presence of at least one male worm) | Yes (male potential 1000) | Assumed | Yes (male potential 1000) | Assumed |
| Mating cycle (number of months a female can produce mf with one insemination) | 1 | Assumed | 1 | Assumed |
| Variation in worm contribution to mf count (dispersal) | No variation | Assumed | No variation | Assumed |
| **Uptake of infection by the vector** |  |  |  |  |
| Functional relationship | $L3=a(1-\exp\left( -(bM \right)^{c}))$  a = 1.666  b = 0.026  c = 1.514 | Previously estimated by fitting model to data [7] | ${L3}_{i}\left( t \right)= c+\frac{a{\cdot m}_{i}(t)}{1+a/b\cdot m_{i}(t)}$  a = 0.089 (scale parameter, denoting the mf load (*m_i_*) at which changes occur)  b = 6.6 (maximum L3 load)  c = 0 (intercept) | [11] |
| Transmission probability (v), fraction of the L3 larvae, resulting from a single blood meal, that is released by a mosquito | 0.1 | Fixed, as in [3] | 0.1 | Fixed, as in [3] |
| **Surveillance** |  |  |  |  |
| Timing of surveys | Yearly, at the start of each year, i.e. just before an MDA round |  | Yearly, at the start of each year, i.e. just before an MDA round |  |
| Volume of blood examined for mf | 60 μL | Assumed | 60 μL | Assumed |
| Variability in observed number of mf in one 20 μl blood smear | Negative binomial distribution with k=0.33 | Previously estimated for 20 μL blood by fitting to data, [3] | Negative binomial distribution with k=0.345 | Previously estimated for 20 μL blood by fitting to data [3] |
| Variation between worms in their contribution to measured mf count (dispersal factor) | Constant (no variation) | Assumed | Constant (no variation) | Assumed |
| **Morbidity & excess mortality due to disease** | Not considered |  | Not considered |  |
| **MDA and vector control** |  |  |  |  |
| Timing and coverage of treatment |  | |  |  |
| Fraction of the population never participating in treatment (= proportion systematic non-compliers) |  | |  |  |
| Minimum age for treatment |  | |  |  |
| Relative compliance by age and sex | 0 below the minimum age for treatment and 1 at and above the minimum age for treatment .  No differences between sexes | Assumed | 0 below the minimum age for treatment and 1 at and above the minimum age for treatment .  No differences between sexes | Assumed |
| Vector control | Not considered simulated scenarios |  | Not considered simulated scenarios |  |
| Interventions modifying individual’s exposure to mosquito bites (exposure / contribution interventions) | Not considered simulated scenarios |  | Not considered simulated scenarios |  |

The relative monthly biting rate (mbr) and the shape (=rate) parameter (k) of the Gamma distribution describing exposure heterogeneity in the simulated population were varied between runs, in order to generate simulations across a wide range of mf prevalences at baseline, measured in the population aged 5 and above. For Africa, we simulated settings with mean prevalence of 10%, 20% and 30%; for India, we simulated settings with mf prevalences of 5% and 10% in the specified age groups, allowing for some variation around this mean (2.5%-point below and above the targeted mf prevalence, with simulated prevalences evenly distributed within this range).

For Africa, the shape parameter of the gamma distribution was allowed to vary between 0.13 and 0.95, and the monthly biting rate was allowed to vary between 150 and 1600). For India, the shape parameter of the gamma distribution was allowed to vary between 0.1 and 1.4, and the monthly biting rate was allowed to vary between 300 and 3000), informed by previous simulations with these model variants. We assumed no importation of infection from surrounding areas (i.e. the external force of infection = 0). Selected parameter values are shown in Figure S5 (Africa) and Figure S6 (India).

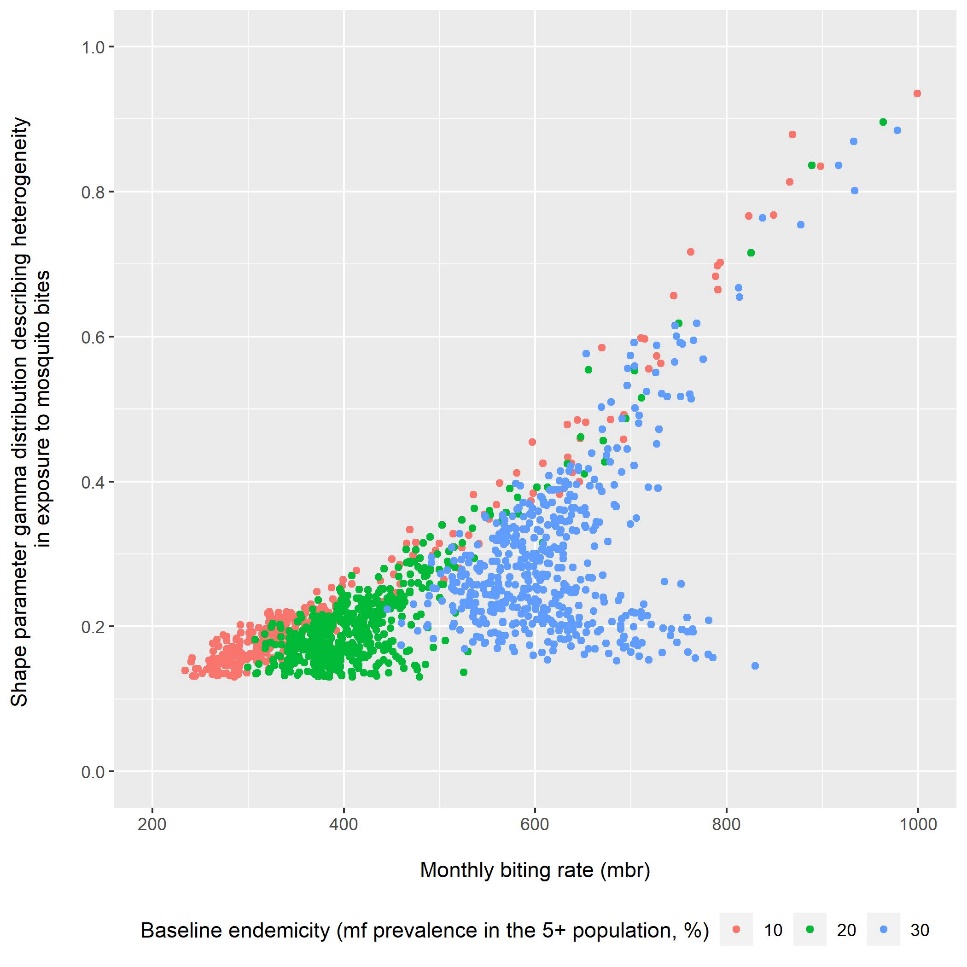

**Figure S5:** Parameter-combinations considered to simulate Mf prevalence of 10%, 20%, 30% in the population aged 5 years and above for Africa.

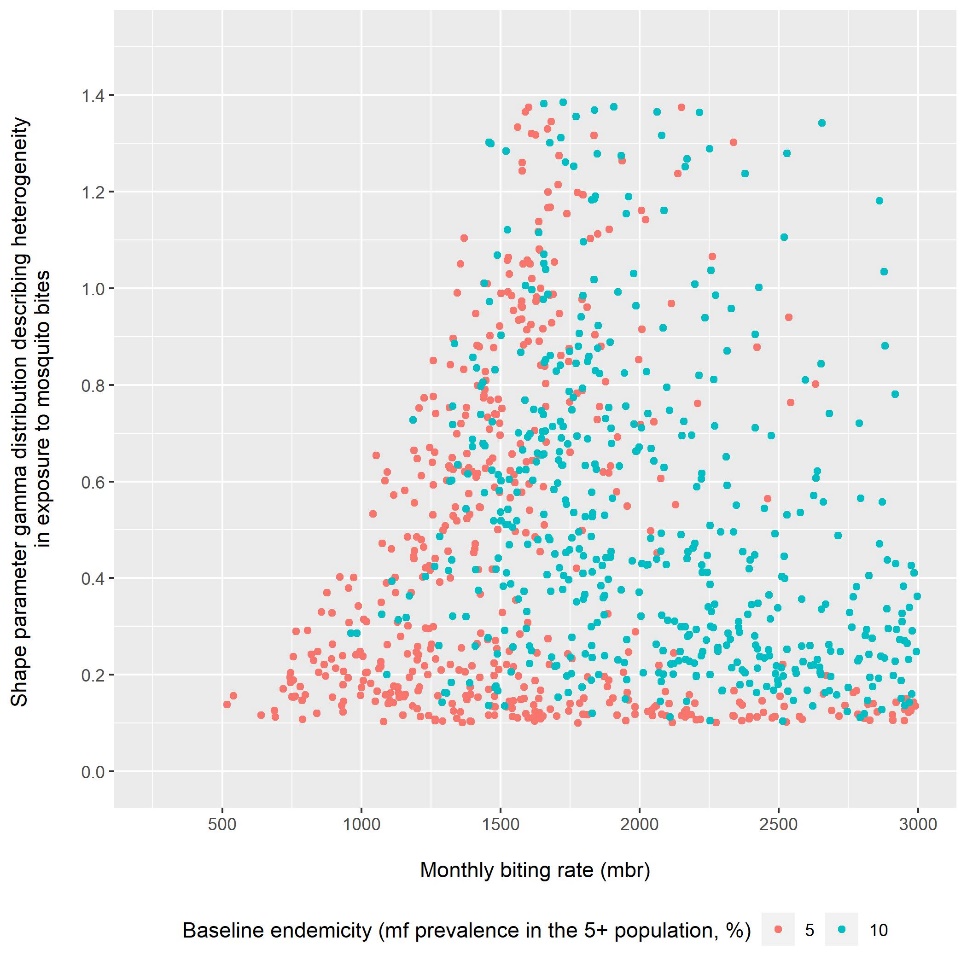

**Figure S6:** Parameter-combinations considered to simulate Mf prevalence of 5% or 10% in the population aged 5 years and above for India.

### 3.3. Modelling never treatment

Scenarios are defined in terms of the achieved coverage among eligible age groups and the proportion never treated among people eligible in all rounds. For LYMFASIM, however, requires input of the coverage achieved in the total population (including ineligible age groups) and the fraction of people that will never participate in MDA. The association between those parameters depend on the age structure of the simulated population and the minimum age for treatment. The tables below give the input specification corresponding to the scenarios to be simulated.

**Table S8:** LYMFASIM inputs on coverage and fraction that will never be treated used to simulate given combination of coverage in eligible age groups and the fraction that was never treated after 5 treatment rounds, for the Africa model and drug combinations provided people aged 2 and above.

| **Coverage in eligible age group** | **Corresponding coverage in total population** | **Fraction of the population never treated required reach the given fraction never treated after 5 rounds** | | | | | | | |
| --- | --- | --- | --- | --- | --- | --- | --- | --- | --- |
|  |  | **0** | **0.05** | **0.1** | **0.15** | **0.2** | **0.25** | **0.3** | **0.35** |
| **0.65** | 0.603 |  | 0 | 0.068 | 0.135 | 0.194 | 0.249 | 0.3 | 0.35 |
| **0.7** | 0.649 |  | 0.023 | 0.087 | 0.145 | 0.199 | 0.25 | 0.3 |  |
| **0.75** | 0.6968 |  | 0.039 | 0.096 | 0.149 | 0.2 | 0.25 |  |  |
| **0.8** | 0.742 | 0 | 0.047 | 0.099 | 0.15 | 0.2 |  |  |  |
| **0.85** | 0.789 | 0 | 0.049 | 0.1 | 0.15 |  |  |  |  |
| **0.9** | 0.835 | 0 | 0.05 | 0.1 |  |  |  |  |  |
| **0.95** | 0.881 | 0 | 0.05 |  |  |  |  |  |  |

**Table S9:** LYMFASIM inputs on coverage and fraction that will never be treated used to simulate given combination of coverage in eligible age groups and the fraction that was never treated after 5 treatment rounds, for the Africa model and drug combinations provided people aged 5 and above.

| **Coverage in eligible age group** | **Corresponding coverage in total population** | **Fraction of the population never treated required reach the given fraction never treated after 5 rounds** | | | | | | | |
| --- | --- | --- | --- | --- | --- | --- | --- | --- | --- |
|  |  | **0** | **0.05** | **0.1** | **0.15** | **0.2** | **0.25** | **0.3** | **0.35** |
| **0.65** | 0.543 |  | 0 | 0.067 | 0.134 | 0.194 | 0.249 | 0.3 | 0.35 |
| **0.7** | 0.585 |  | 0.022 | 0.087 | 0.145 | 0.199 | 0.25 | 0.3 |  |
| **0.75** | 0.627 |  | 0.039 | 0.096 | 0.149 | 0.2 | 0.25 |  |  |
| **0.8** | 0.668 | 0 | 0.047 | 0.099 | 0.15 | 0.2 |  |  |  |
| **0.85** | 0.710 | 0 | 0.049 | 0.1 | 0.15 |  |  |  |  |
| **0.9** | 0.752 | 0 | 0.05 | 0.1 |  |  |  |  |  |
| **0.95** | 0.794 | 0 | 0.05 |  |  |  |  |  |  |

**Table S10:** LYMFASIM inputs on coverage and fraction that will never be treated used to simulate given combination of coverage in eligible age groups and the fraction that was never treated after 5 treatment rounds, for the India model and drug combinations provided people aged 2 and above.

| **Coverage in eligible age group** | **Corresponding coverage in total population** | **Fraction of the population never treated required reach the given fraction never treated after 5 rounds** | | | | | | | |
| --- | --- | --- | --- | --- | --- | --- | --- | --- | --- |
|  |  | **0** | **0.05** | **0.1** | **0.15** | **0.2** | **0.25** | **0.3** | **0.35** |
| **0.65** | 0.612452347 |  | 0 | 0.068 | 0.135 | 0.194 | 0.249 | 0.3 | 0.35 |
| **0.7** | 0.659564066 |  | 0.023 | 0.088 | 0.145 | 0.199 | 0.25 | 0.3 |  |
| **0.75** | 0.706675784 |  | 0.039 | 0.096 | 0.149 | 0.2 | 0.25 |  |  |
| **0.8** | 0.753787503 | 0 | 0.047 | 0.099 | 0.15 | 0.2 |  |  |  |
| **0.85** | 0.800899222 | 0 | 0.049 | 0.1 | 0.15 |  |  |  |  |
| **0.9** | 0.848010941 | 0 | 0.05 | 0.1 |  |  |  |  |  |
| **0.95** | 0.89512266 | 0 | 0.05 |  |  |  |  |  |  |

### 3.4. Model version and availability

Model version used in this study: wormsim version 2.58Ap59.

Programme and source code availability: LYMFASIM was originally developed as a standalone computer programme [1], but is now incorporated as a disease-specific variant within WORMSIM, a generalized framework for modelling transmission and control of helminth infections in humans. A formal description of WORMSIM has been provided elsewhere for version v2.58Ap9 [2]. The programme and source code are available at gitlab:

<https://gitlab.com/erasmusmc-public-health/wormsim.previous.versions>

### 3.5. References

1. Plaisier AP, Subramanian S, Das PK, Souza W, Lapa T, et al. (1998) The LYMFASIM simulation program for modeling lymphatic filariasis and its control. Methods of Information in Medicine 37: 97-108.

2. Coffeng LE, Bakker R, Montresor A, de Vlas SJ (2015) Feasibility of controlling hookworm infection through preventive chemotherapy: a simulation study using the individual-based WORMSIM modelling framework. Parasit Vectors 8: 541.

3. Subramanian S, Stolk WA, Ramaiah KD, Plaisier AP, Krishnamoorthy K, et al. (2004) The dynamics of *Wuchereria bancrofti* infection: a model-based analysis of longitudinal data from Pondicherry, India. Parasitology 128: 467-482.

4. Stolk WA, ten Bosch QA, de Vlas SJ, Fischer PU, Weil GJ, et al. (2013) Modeling the impact and costs of semiannual mass drug administration for accelerated elimination of lymphatic filariasis. PLoS Negl Trop Dis 7: e1984.

5. Smith ME, Singh BK, Irvine MA, Stolk WA, Subramanian S, et al. (2017) Predicting lymphatic filariasis transmission and elimination dynamics using a multi-model ensemble framework. Epidemics 18: 16-28.

6. Stolk WA, Prada JM, Smith ME, Kontoroupis P, de Vos AS, et al. (2018) Are Alternative Strategies Required to Accelerate the Global Elimination of Lymphatic Filariasis? Insights From Mathematical Models. Clin Infect Dis 66: S260-S266.

7. Stolk WA, de Vlas SJ, Borsboom GJ, Habbema JD (2008) LYMFASIM, a simulation model for predicting the impact of lymphatic filariasis control: quantification for African villages. Parasitology 135: 1583-1598.

8. Prada JM, Davis EL, Touloupou P, Stolk WA, Kontoroupis P, et al. (2020) Elimination or Resurgence: Modelling Lymphatic Filariasis After Reaching the 1% Microfilaremia Prevalence Threshold. J Infect Dis 221: S503-S509.

9. World Health Organization (1992) Lymphatic filariasis: the disease and its control. Fifth report of the WHO Expert Committee on Filariasis. World Health Organ Tech Rep Ser 821: 1-71.

10. Plaisier AP, Cao WC, van Oortmarssen GJ, Habbema JD (1999) Efficacy of ivermectin in the treatment of *Wuchereria bancrofti* infection: a model-based analysis of trial results. Parasitology 119: 385-394.

11. Subramanian S, Krishnamoorthy K, Ramaiah KD, Habbema JDF, Das PK, et al. (1998) The relationship between microfilarial load in the human host and uptake and development of *Wuchereria bancrofti* microfilariae by *Culex quinquefasciatus*: a study under natural conditions. Parasitology 116: 243-255.

### 4. PRIME-NTD table

**Table S10:** The Policy-Relevant Items for Reporting Models in Epidemiology of Neglected Tropical Diseases (PRIME-NTD).

| **Principle** | **What has been done to satisfy the principle?** | **Where in the manuscript is this described?** |
| --- | --- | --- |
| **Stakeholder engagement** | Reference to WHO guideline/roadmap. Work has been presented at the following WHO / NTD Modelling Consortium meeting: “Accelerating progress towards the 2030 NTD targets: How can quantitative modelling support programmatic decisions?” | Introduction (the reference to WHO guideline/roadmap). |
| **Complete model documentation** | Transmission models are described in the manuscript. | Methods and Supplementary Information. |
| **Complete description of data used** | Parameters used are described in the manuscript. | Table 1, Table S5, Table S7-S10 |
| **Communicating uncertainty** | We have considered two different transmission models and discussed other factors, not included in this analysis, that can influence the effect of NT on achieving elimination. | Methods and Discussion sections. |
| **Testable model outcomes** | The model outcomes can be tested by collecting data on never treatment. | Discussion section. |
